# Ultra-processed food consumption and risk of lung cancer: Results from a Prospective Study

**DOI:** 10.1101/2024.09.27.24314515

**Authors:** Tefera Chane Mekonnen, Yohannes Adama Melaka, Zumin Shi, Tiffany K Gill

## Abstract

**Background and Aims:** There is limited evidence on the link between ultra-processed food (UPF) intake and the risk of lung cancer (LC). This study examined the association between UPF and LC risk using data from the Prostate, Lung, Colorectal, and Ovarian (PLCO) cancer trial.

**Methods:** This study involved PLCO participants (n = 96,607, aged ≥ 55 years) who were followed between 1998 and 2009. Food items were categorized based on the NOVA classification. Cox regression models with inverse probability of censoring weighting (IPCW) were utilized to estimate the association between UPF intake and LC risk. The joint effect of UPF and diabetes was explored using additive hazard models to calculate the additional number of LC cases.

**Results:** During a median follow-up period of 9.4 years, 1,596 incident LC cases were identified. UPF consumption (in %gram/day) showed no significant association with the overall risk of LC. However, adults with diabetes in the highest quintile of UPF intake had a significantly higher risk of LC (HR = 2.44; 95% CI: 1.27, 4.67) compared to participants without diabetes. A small excess risk due to the interaction between UPF and diabetes (0.13; 95% CI −0.32, 0.58) was observed, resulting in an additional 201 cases of LC per 10^5^ person-years (95% CI: 70, 332) attributed to the highest UPF intake and diabetes interaction. Furthermore, a 10% increment in UPF intake (%kcal/day) increased the risk of LC by 32%.

**Conclusions:** While UPF, in terms of weight contribution, is associated with a higher risk of LC in participants with diabetes, UPF (in %kcal/day), is associated with an increased risk of LC in all participants. Lowering UPF intake may help reduce the risk of LC in both diabetic patients and the general population.

## Introduction

Lung cancer (LC) is one of the most common and deadliest types of cancer worldwide and its incidence has been increasing in many countries over the past few decades [1]. It is estimated that approximately 2.3 million people are diagnosed with LC each year, over 2 million people die from the disease annually and LC accounts for 45.9 million disability-adjusted life years (DALYs) in both sexes [2, 3]. LC is the leading cause of cancer death, accounting for 23% of all cancer deaths in America [4]. While smoking is the primary cause of LC, there is growing concern that dietary factors may also play a role in the development of the disease [5, 6]. Ultra-processed food (UPF) is typically highly processed and contain a large number of additives, such as artificial flavours, colours, and preservatives. The consumption of UPF in the modern diet has increased dramatically in recent years, and this has raised concerns about their potential impact on health [7].

There is limited scientific evidence directly linking the consumption of UPF to an increased risk of LC. However, there is a growing body of research suggesting that a diet high in UPF may increase the risk of other types of cancer, as well as other chronic diseases [8–10]. A recent (2023) systematic review and meta-analysis, which did not include LC, found that a higher proportion of UPF intake increases the risk of overall cancer by 13%, breast cancer by 11%, colorectal cancer by 30% and pancreatic cancer by 49% [11]. Consistent epidemiological findings from population-based studies (UK biobank, the European Prospective Investigation into Cancer and Nutrition (EPIC) study, the French NutriNet-Sante cohort and US-based cohorts) have reported a link between UPF consumption and overall cancer, ovarian, colorectal, breast and pancreatic cancer [12–17]. Only two of these studies evaluated the association of UPF and LC. These two studies have investigated the association between UPF and LC using UK biobank and EPIC study and shown no association. One study found that a diet high in UPF was associated with an increased risk of all types of cancer, including LC. However, the study did not specifically examine the relationship between ultra-processed food consumption and LC risk [18].

While the relationship between UPF consumption and LC risk is not yet fully understood, there is some evidence to suggest that a diet high in ultra-processed foods may be linked to an increased risk of LC [19–24]. Examining the association and understanding the potential link between UPF consumption and LC risk is important for developing effective strategies to prevent this disease. Hence, the current study was aimed to investigate the association between proportion of UPF intake and LC among older adults in United States of America (USA).

## Methods

### Study Population and Study Design

The study used data from the Prostate, Lung, Ovarian and Colorectal Cancer trial (PLCO), a cancer screening trial in 10 study centers across the USA. The trial was designed to evaluate whether screening examinations reduces cancer mortality by randomly allocating approximately 155,000 older adults to the control arm (usual care) or to the intervention arm (screening examination). The randomization began in 1993. Eligible study subjects were aged between 55 to 74 years at enrolment and free from prostate, lung, colorectal and ovarian cancer, with additional criteria documented elsewhere [25]. Initial information on demographic, lifestyle, medical history, family history and medication used was collected through the baseline questionnaire (BQ). The dietary history questionnaire (DHQ) was administered to both arms commencing in 1998 and a total of 113,000 (77%) participants provided a complete response with a 3-year median time to the study. Study subjects were monitored approximately for 12 years to obtain data related to the diagnosis of LC.

Overall, 101,732 participants were eligible for dietary history and LC analysis (53,155 study participants were excluded due to invalid BQ and DHQ). After examining the distribution of total energy, we further excluded 5,125 participants who were in the bottom and top 1% of energy intakes, with 96,607 participants remaining in the final analysis (**Supplementary** Fig 1). Ethics approval for the trial was obtained from the US National Cancer Institute (NCI) Review Boards and written informed consent was obtained from each participant. The data for dietary history and lung cancer analysis were obtained using the Cancer Data Access System (CDAS) after obtaining approval from NCI (project registration: PLCO-982).

### Data Collection and UPF Assessment

Baseline data on demographic characteristics (age, sex, race, marital status, education, occupation, family income and study arm), types and date of diagnosis for cancer, treatment, self-reported medical history, personal lifestyle features (smoking, alcohol drinking, and body-mass index (BMI)), family history of lung cancer, medication use, physical activity and dietary habits were obtained from the BQ, DHQ, supplementary questionnaire (SQ) and brief survey questionnaire (BSQ). Dietary data were collected using a self-administered food frequency questionnaire (FFQ), a validated assessment tool which provides better nutrient estimate[26]. All food items in the DHQ were grouped into one of four categories from the NOVA classification, with an emphasis on UPF as described previously by Monteiro [27]. In case of uncertainty regarding which category the food or beverage belongs to, consensus was reached among the researchers (**Supplementary Table 1**).

From a total of 275 foods or beverages, 145 were categorized as UPF. UPF in this study includes carbonated drinks, savory packaged snacks; ice cream, chocolate, confectionery; breads and buns; margarines and spreads; cookies, pastries, and cakes; breakfast cereals, cereal and energy bars; flavoured milk drinks; cocoa drinks; sweet desserts made from fruit with added sugars, artificial flavours and texturizing agents; cooked seasoned vegetables with ready-made sauces; meat and chicken extracts and instant sauces; health and slimming products such as powdered or fortified meal and dish substitutes; ready to heat products; poultry and fish nuggets and sticks, sausages, burgers, cold cuts, hot dogs, and other reconstituted meat products, and instant soups, and noodles. These food items were further regrouped into sugary drinks; processed meats; milk dessert, yogurt, and soy products; cookies, pies, and pastries; margarine and dressings; sweets and other condiments; salty snacks; quick bread; and others (**Supplementary Table 1**).

UPF was measured by calculating the weight ratio of all food items classified as UPF to the total weight of all food items consumed by individuals per day (% grams/day). This weight ratio was chosen instead of an energy ratio because it considers UPF with no or low-calorie content, such as artificially sweetened beverages, as well as non-nutritional factors associated with food processing (such as neoformed contaminants, additives, and alterations to the structure of raw foods)[12, 13, 28]. Additionally, the caloric contribution of UPF was determined by calculating the fraction of energy from UPF to the total energy from all food items consumed per day.

### Outcome Assessment

The outcome of this study was incidence of LC. LC diagnosis was made by self-reported responses from participants through annual follow-up questionnaires. Cases were documented from abnormal x-ray results, death certificates and relative reports. LC diagnosis confirmed from medical record abstraction as per histopathologic type derived from the International Classification of Diseases-Oncology, 2^nd^ edition (ICD-O-2) morphology that includes either non-small cell lung cancer (NSCLC) (n=1,464) or small cell lung cancer (SCLC) (n=242). Lung carcinoid tumour was not considered confirmed LC during the trial and was excluded in this analysis.

### Statistical Analysis

We utilized an inverse probability of censoring weighting (IPCW) estimator to reduce bias caused by censoring, specifically dependent censoring [29, 30]. This method has been employed in right-truncated data [31] and can address the issue of censored subjects by assigning greater weight to subjects with similar characteristics who are not censored [32]. By making the positivity assumption, this approach enables us to consistently estimate the effects of covariates and avoids the need to estimate baseline hazards [31].

We applied Cox regression models with IPCW to estimate the hazard ratio (HR) and 95% confidence interval (CI) for the association between proportion of UPF consumption and LC risk. The follow-up time was defined as the time between completion of DHQ and date of diagnosis of LC, death, drop-out, or the end of the study up to 31 December 2009. We built five models in incremental steps by adjusting for potential confounders, selected from literature and prior knowledge [12, 13, 28, 33]. These covariates considered for the model adjustment were age (continuous), sex (men, women), marital status (married, widowed, divorced, separated/never married), education (up to grade 12 completion, post high school and some college, and under and postgraduate), ethnicity (Hispanic or non-Hispanic), study arm (control, intervention), BMI (<18.5, 18.5-24.99, 25-29.99, ≥30+), cigarette smoking status (never, current, former), alcohol intake(g/day), total energy (kcal/day), aspirin use (yes, no), history of baseline comorbidity (yes, no), and family history of LC (yes, no). Models were also adjusted for average family income (<$50,000, $50,000-$99,000 and ≥ $100,000) and physical activity measured in total time spent (in minutes) during each session doing moderate-to-strenuous exercise, as recorded in the self-reported SQ. Since cigarette smoking is a well-established risk factor for LC, we further refined the models by considering the intensity and duration of smoking for both current and former smokers. Smokers were classified into the following groups: never smokers, current smokers consuming 1-10 cigarettes per day, current smokers consuming 11-20 cigarettes per day, current smokers consuming 21 or more cigarettes per day, former smokers who quit within the past 10 years, former smokers who quit 11-20 years ago, and former smokers who quit more than 20 years ago [13, 34]. We also consider cigar and pipe smoking in the analyses.

Cox regression with restricted cubic splines was performed to examine the relationship between per 10% increment in UPF intake and LC risk with three knots (10^th^, 50^th^, and 90^th^), selected based on Akaike Information Criterion (AIC). In addition to using IPCW, we conducted competing risk regression analyses to consider the impact of deaths that occurred before the diagnosis of LC on the observed relationship between the quintile of UPF consumption and the risk of LC. Subgroup analysis was carried out by stratifying the analysis by sex, age group, race, cigarette smoking, BMI, family history of LC, history of hypertension, diabetes, chronic bronchitis, and emphysema to determine whether associations were modified by multiplicative interactions of covariates and avoid potentially misleading subgroup differences.

Effect modification was assessed using Cox regression to determine the relative excess risk due to interaction (RERI: RR11−RR10−RR01+1), attributable proportion (AP: RERI/RR11), and synergy index (SI: (RR11−1)/(RR10+RR01−1)) on an additive scale, with 95% confidence intervals [35]. RR11 represents the relative risk (RR) in the groups with high UPF exposure and diabetes exposure, RR10 represents the RR in the groups with high UPF intake and no diabetes, and RR01 represents the RR in the groups with low UPF intake and diabetes. Interactions between UPF consumption and diabetes status were analysed using the surrogate measures mentioned earlier. A value of zero for RERI and AP indicates no additive interaction. A positive value suggests a super-additive interaction, while a negative value indicates a sub-additive interaction [36, 37]. A value of 1 for SI implies no interaction or exactly additivity; SI > 1 indicates positive interaction or more than additivity; SI < 1 suggests negative interaction or less than additivity [37]. The study also examined the interaction between diabetes status (yes/no) and the quintile of UPF exposure to compare the outcomes of both types of exposure.

In Cox models, deviations from additivity can be assessed using the surrogate measures of additive interaction obtained from the multiplicative models mentioned above. However, these measures may sometimes be counter-intuitive and invalid. An alternative approach is to use additive hazard models, which directly estimates the absolute magnitude of the deviation from additivity [38, 39]. Therefore, we used additive hazard models to estimate the number of additional lung cancer incidents per 100,000 person-years of observation. We considered the quintiles or binary categories of UPF intake (low, below the mean (<31.2%), and high, ≥ the mean (≥31.2%)), diabetes status, and their interaction using the R package “timereg” [38]. Finally, to check the stability of the findings, we performed the following sensitivity analyses: a) multiple imputation for covariates with missing values including physical activity (23,002; 24%) and family income (26,629; 28%); b) excluding participants with history of hypertension, diabetes, chronic bronchitis, emphysema and obese at baseline; c) excluding LC cases that occurred during the first five years of follow-up; d) Adjusting for processed meat, artificial sweetening beverages, fruits and vegetables, fish consumption and nutrients such as fibers, omega-3, total fat, protein, carbohydrates, and sodium; e)comparing the findings with the proportion of UPF (%kcal) of the total energy; g) considering histopathological subtypes of LC (NSCLC and SCLC) and h) removing putative confounders (smoking, alcohol intake, and BMI) of lung cancer in the final model to see their impact on the observed association between UPF intake and LC risk. All statistical analyses were performed using R-software version 4.2.3 and Stata version 18 (College Station, TX, USA). **Results**

### Baseline Characteristics of Study Participants

Among the total participants, 50,803(52.6%) were female and the mean (SD) age was 65.58(5.74) years at baseline for all participants. The proportion of women was consistently lower across the quintiles UPF consumption from 66.5% to 43.5% compared to men. A slightly higher pattern of UPF consumption was observed in younger adults (mean age of 64.6 vs 66 years). In addition, compared to participants in the first quintile of UPF consumption, the number of participants in the fifth quintile was higher among those classified as obese (18.1% vs 27.8%), current cigarette smokers (6.3% vs 12.8%), current alcohol drinkers (13.3% vs 18.7%), history of diabetes (6.2% vs 7.8%), and hypertensive (30.1% vs 34.6 (**Table 1**).

**Table 1:**
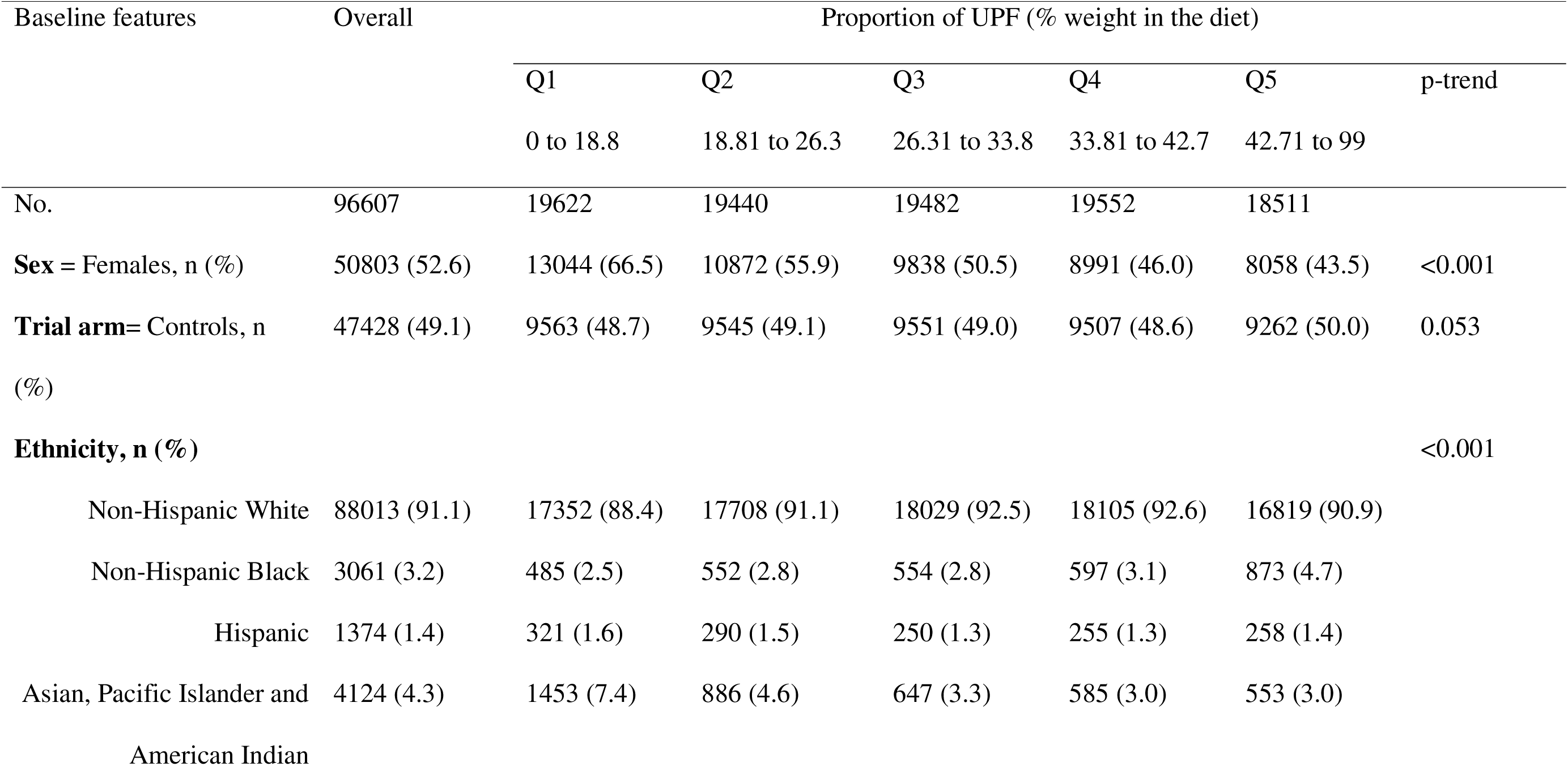
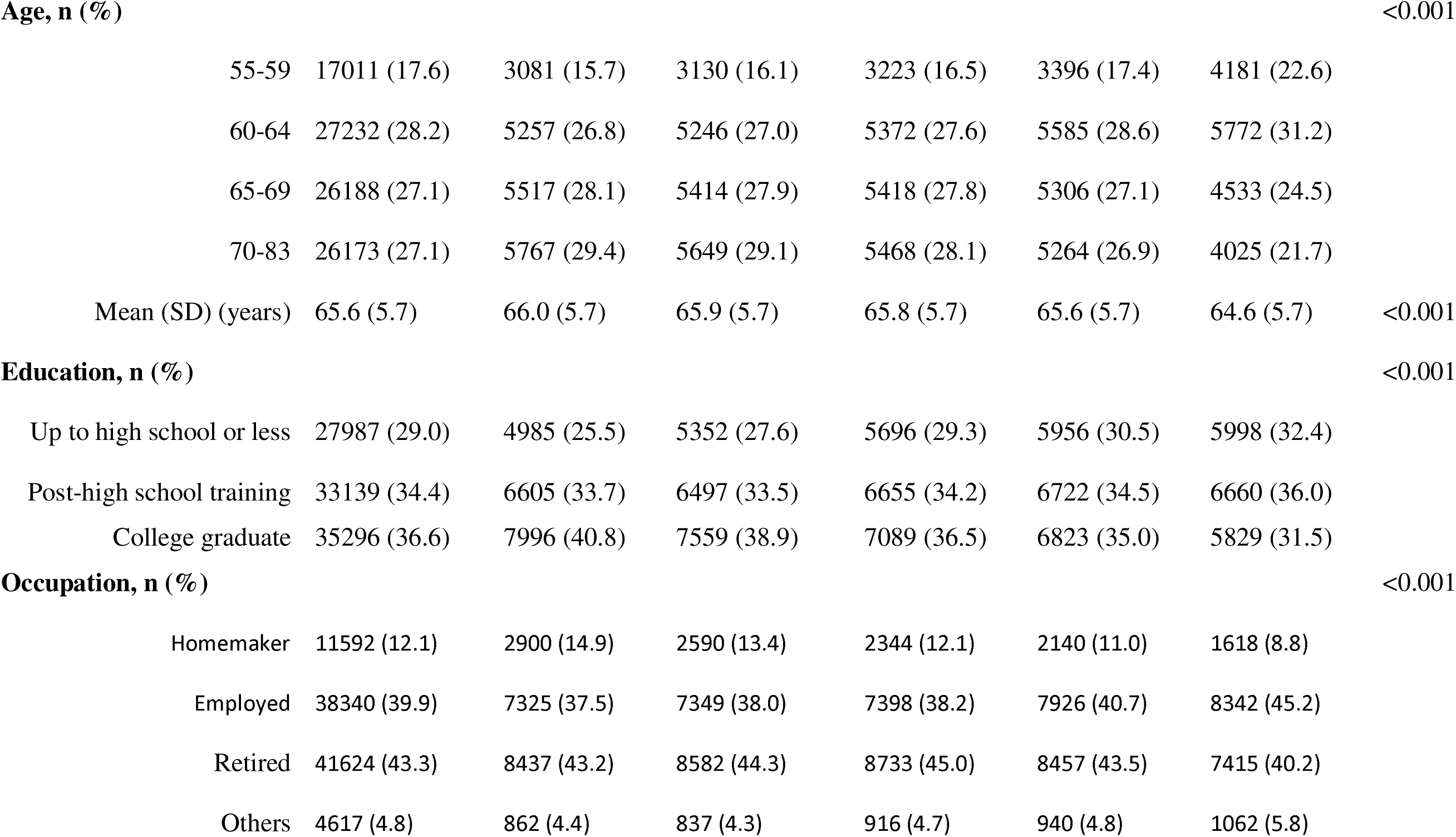
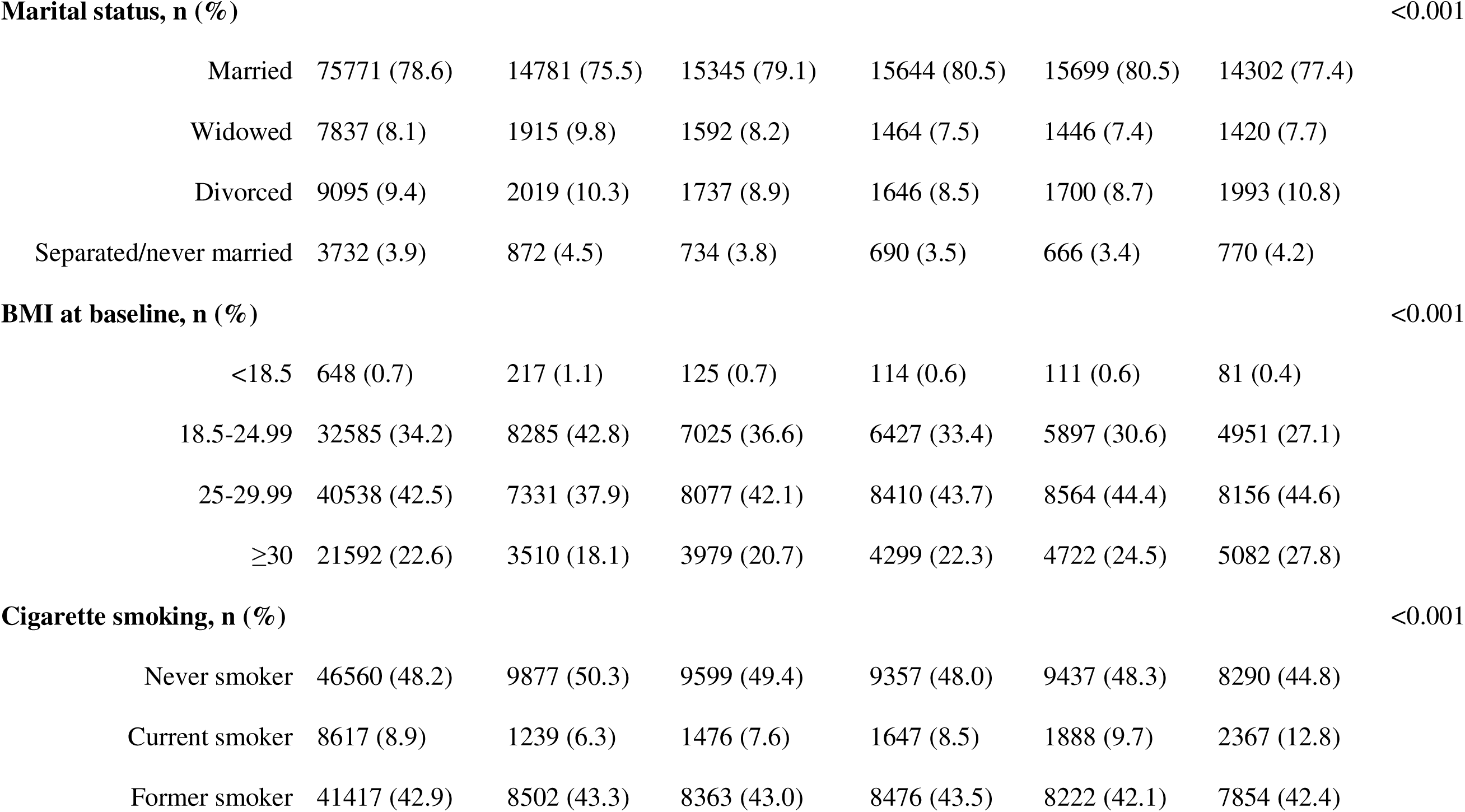
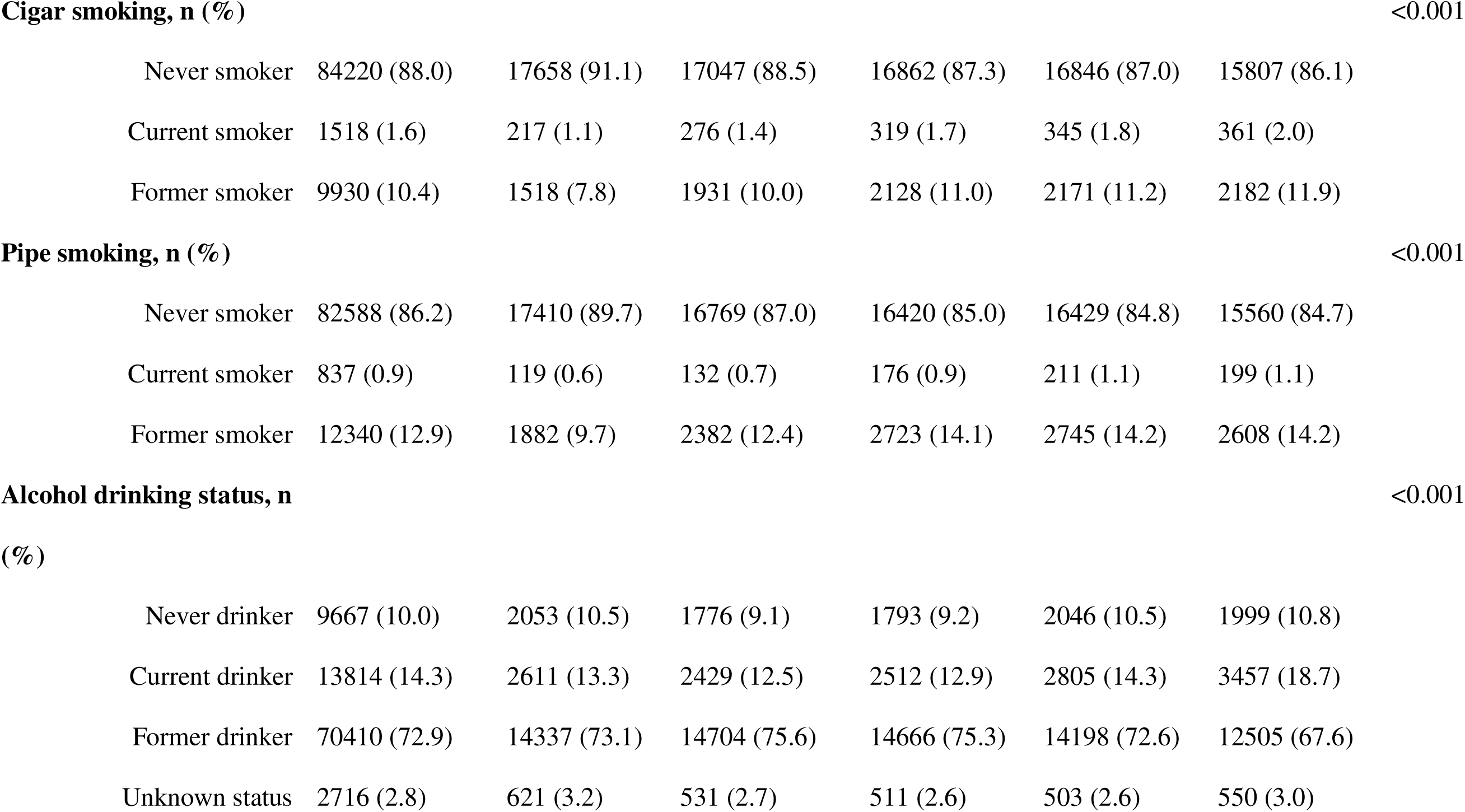
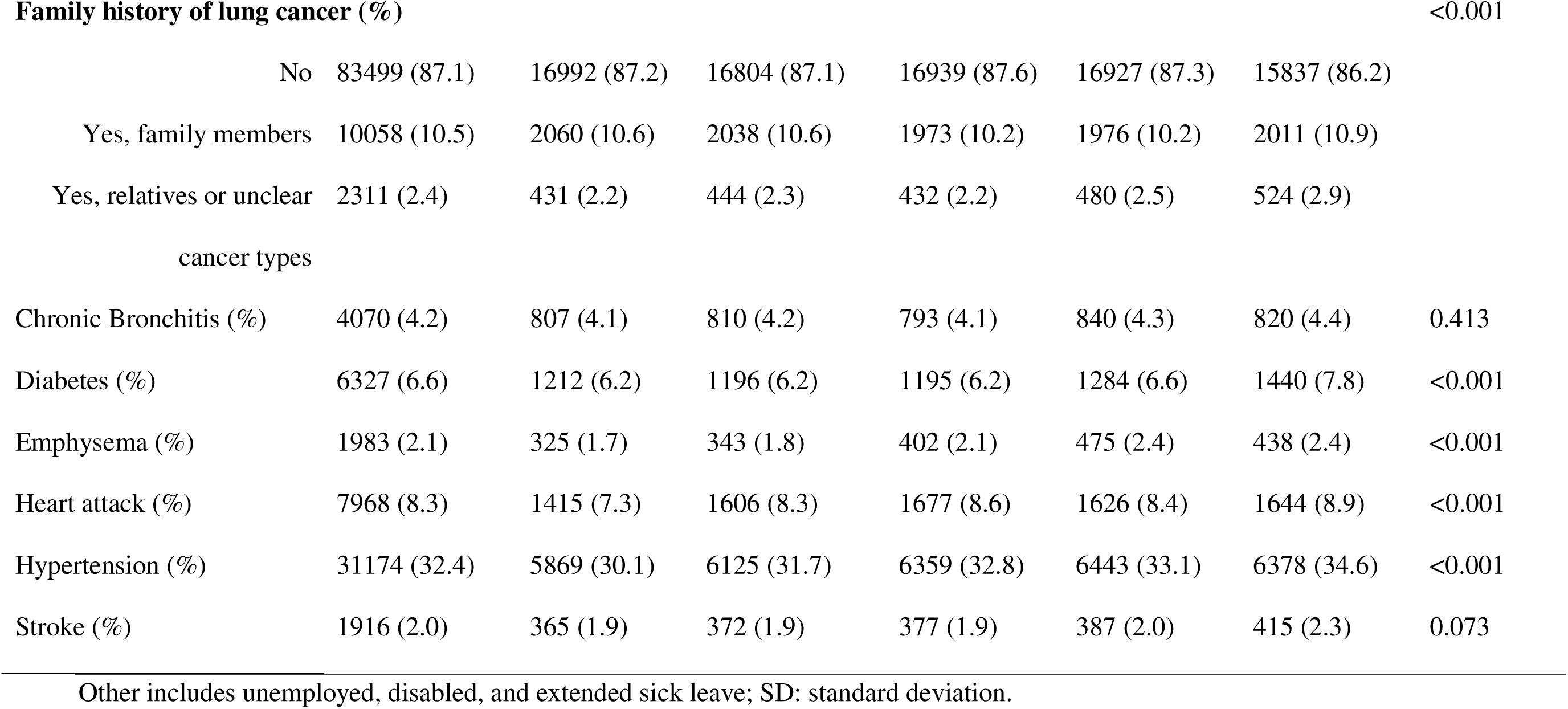
Distribution of baseline characteristics of older adults enrolled in the PLCO trial across quintiles of UPF consumption in USA (n=96607).

The average (SD) proportion of UPF consumption in the total diet (gm/day) was 31.2% (14.0%) (**Supplementary** Fig 2) and the mean percentage of UPF consumption (%kcal) was 37.13% (**Supplementary** Fig 3). Sugary drinks, milk dessert and processed meats contributed higher proportion to the total diet in the study participants (**Fig 1a**). The mean proportion of UPF contribution was relatively higher in men than women 33.45% vs 29.2%. The predominant UPF groups consumed by both men and women were sugary drinks, milk dessert, yogurts, and soy products, processed (white and red) meat, cakes, cookies, pies and pastries, quick breads, and sugar, condiments, and sweets (**Fig 1b, c**).

### UPF Consumption and Lung Cancer Incidence

A total of 1,596 incident cases of LC occurred during a median [IQR] follow up of 9.4[8.02, 10.11) years and mean (SD) follow-up time 8.82(1.93) years. Over a total 853,968.16 person-years, the overall incidence rate of LC was found to be 18.8 (95% CI: 17.7, 19.6) cases per 10,000 individuals. The incidence rate is higher in study subjects in the highest quintile of UPF consumption compared to lowest (21 vs 18 per 10,000 observations) (**Supplementary Table 2**).

Following adjustment for all covariates, the risk of LC incidence had no significant association with a higher consumption of UPF (HR = 0.98; 95% CI: 0.83,1.15, p-for trend =0.32). Results from competing risk regression did not indicate significant association (HR = 0.95; 95% CI: 0.81,1.12). Furthermore, restricted cubic spline analysis did not demonstrate a significant relationship between UPF consumption and LC incidence (HR = 0.98; 95% CI: 0.85,1.13; p-nonlinear =0.44) (**Table 2**, **Fig 2**).

**Table 2:** Cox proportional hazard model with time-varying covariate for the association between proportion of UPF consumption (% gm/day and risk of lung cancer incidence among older population of PLCO trial (Complete cases = 91,453).

| Explanatory variables | Proportion of UPF consumption in the diet |  |  |  |  |  | UPF as continuous exposure |  |
| --- | --- | --- | --- | --- | --- | --- | --- | --- |
|  | HR (95% CI) |  |  |  |  |  | (10% increment) |  |
|  | Q1 | Q2 | Q3 | Q4 | Q5 | p-trend | HR (95% CI) | p-nonlinear |
| Cases | 282 | 280 | 311 | 321 | 323 |  | 1517 |  |
| Person-years | 161583.5 | 163458.8 | 164101.1 | 164313.0 | 153729.7 |  | 807186.2 |  |
| Cumulative Incidence<sup>a</sup> | 17.5(15.5, 19.6) | 17.1(15.2, 19.3) | 18.9(16.9,21.2) | 19.5(17.5, 21.8) | 21.0(18.8, 23.4) |  | 18.8(17.9,19.8) |  |
| Model 1 | Ref | 0.96(0.81,1.12) | 1.06(0.91,1.24) | 1.10(0.95, 1.28) | 1.20(1.02,1.39) * | 0.07 | 1.12(0.97, 1.29) * | 0.57 |
| Model 2 | Ref | 0.92(0.78,1.08) | 1.01(0.86,1.18) | 1.05(0.89, 1.22) | 1.19(1.01,1.39) * | 0.08 | 1.09(0.95, 1.26) *** | 0.08 |
| Model 3 | Ref | 0.91(0.77,1.06) | 0.95(0.81,1.12) | 0.95(0.81, 1.11) | 0.98(0.83,1.15) | 0.33 | 0.99(0.87, 1.15) | 0.36 |
| Model 4 | Ref | 0.91(0.77,1.07) | 0.96(0.81,1.12) | 0.94(0.81,1.11) | 0.95(0.81,1.12) | 0.30 | 0.99(0.86,1.14) | 0.40 |
| Model 5 | Ref | 0.92(0.78,1.08) | 0.98(0.83,1.15) | 0.96(0.82, 1.13) | 0.98(0.83,1.15) | 0.32 | 0.98(0.85, 1.13) | 0.44 |
| Competing risk Regression |  |  |  |  |  |  |  |  |
|  | Ref | 0.92(0.78,1.09) | 0.97(0.82,1.14) | 0.95(0.81,1.12) | 0.95(0.81,1.12) | 0.35 |  |  |

<sup>a</sup> cumulative incidence rate per 10,000 person-years; \* represents significant at p-value <0.05; and \*\*\*significant at p-linear < 0.001.
Model 1: unadjusted model.
Model 2: Adjusted for Age and sex.
Model 3: adjusted for model 2 plus study arm, race, marital status, education, occupation, smoking, BMI, family history of lung cancer and alcohol consumption.
Model 4: Adjusted for model 3 plus chronic comorbidity (diabetes, hypertension, heart attack, stroke, chronic bronchitis, and emphysema).
Model 5: Additionally adjusted for total dietary energy plus model 4 and excluding dietary alcohol consumption.

Subgroup analysis demonstrated that the magnitude of the observed association between risk of LC and UPF consumption stratified by most covariates was unchanged (p-interaction > 0.05). However, when comparing the lowest quintile of UPF consumption to the highest quintile among adults with diabetes, those in the highest quintile had significantly higher risk of LC (HR_quintile5 vs 1_= 2.44; 95%CI: 1.27, 4.67) compared to those without diabetes (HR_quintile5 vs 1_ = 0.92; 95% CI: 0.42, 2; P_interaction_ = 0.002) (**Fig 3**).

A further analysis was conducted to examine the relationship between high and low intake of UPF and diabetes status in relation to the risk of LC. Among individuals without diabetes, those with high UPF intake had a risk of LC of 0.99 (95% CI: 0.89, 1.10) compared to those with low UPF intake. However, for individuals with both high UPF intake and diabetes, the hazard ratio for the risk of LC was 1.17 (95% CI: 0.79, 1.75). There was evidence of a positive interaction on the additive scale (supra-additivity) with RERI = 0.13 (95% CI: −0.32, 0.58), AP = 0.10 (95% CI: −0.22, 0.42), and SI = 1.67 (95% CI: 0.22, 12.46). However, there was no statistically significant interaction between high UPF intake and diabetes on the multiplicative scale (p-value = 0.12). Nevertheless, a significant multiplicative interaction was observed between quintiles of UPF intake and diabetes (p-value = 0.007) (**Table 3**). Using additive hazards regression, it was estimated that 6 (95% CI: −13, 26) cases per 100,000 person-years of LC were attributable to high UPF consumption. For diabetes alone, 14 (95% CI: −50, 77) cases of LC were estimated. Due to the joint effect of high UPF intake and diabetes (both binary exposures), an additional 35 (95% CI: −50, 120) cases of LC were estimated. Among participants with diabetes only, 186 (95% CI: 59, 313) cases per 100,000 person-years of LC were attributable to the highest quintile of UPF exposure. An additional 201 (95% CI: 70, 332) cases of LC were estimated due to the combined interaction between the highest UPF intake and diabetes (**Table 4**).

**Table 3:** Adjusted HRs (95% CI) and additive interaction between UPF consumption and diabetes status for the risk of LC in US (n = 91,453) ^a^.

| Percentage<br>of UPF<br>intake<sup>b</sup> | Subjects without diabetes |  | Subjects with diabetes |  | Diabetic vs non-diabetic in<br>strata of %UPF |  | Interaction (Additive) |  |  |
| --- | --- | --- | --- | --- | --- | --- | --- | --- | --- |
|  | Cases/n | HR (95% CI) | Cases/n | HR (95% CI) | Cases/n | HR (95% CI) | RERI (95%<br>CI) | AP (95% CI) | SI (95% CI) |
| Low | 525/85,512 | Ref | 37/5,941 | Ref | 562/36,563 | 1.01(0.71,1.43) | 0.13(-<br>0.32,0.58) | 0.10(-<br>0.22,0.42) | 1.67(0.22,12.46) |
| High | 879/85,512 | 0.99(0.89,1.11) | 76/5,941 | 1.17(0.79,1.75) | 955/54,890 | 1.22(0.96,1.55) |  |  |  |

AP: Attributable Proportion; CI: Confidence Interval; HR: Hazard Ratio; LC: Lung Cancer; RERI: Relative Excess Risk due to Interaction; SI: Synergy Index; UPF: Ultra-processed Food.
<sup>a</sup> Models were adjusted for age, sex, ethnicity, study arm, marital status, education, occupation, cigarette smoking, BMI, family history of lung cancer, history of hypertension, heart attack, chronic bronchitis, emphysema, alcohol intake and total dietary calory intake.
<sup>b</sup> Percentage of UPF (%gm/day) categorised as high and low based on the mean intake of UPF which was $\geq 31.2\%$ for high and $< 31.2\%$ for low).

**Table 4:**
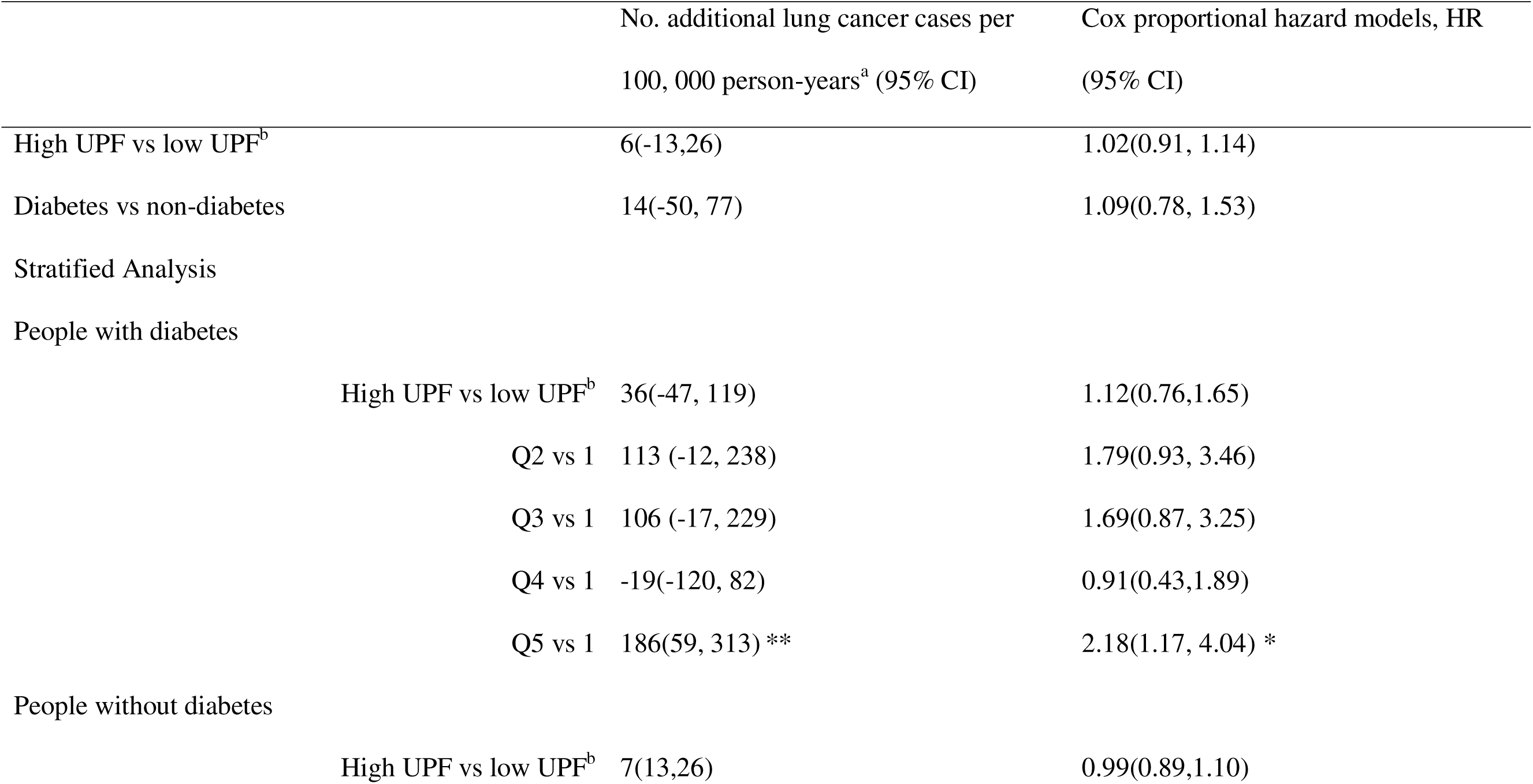
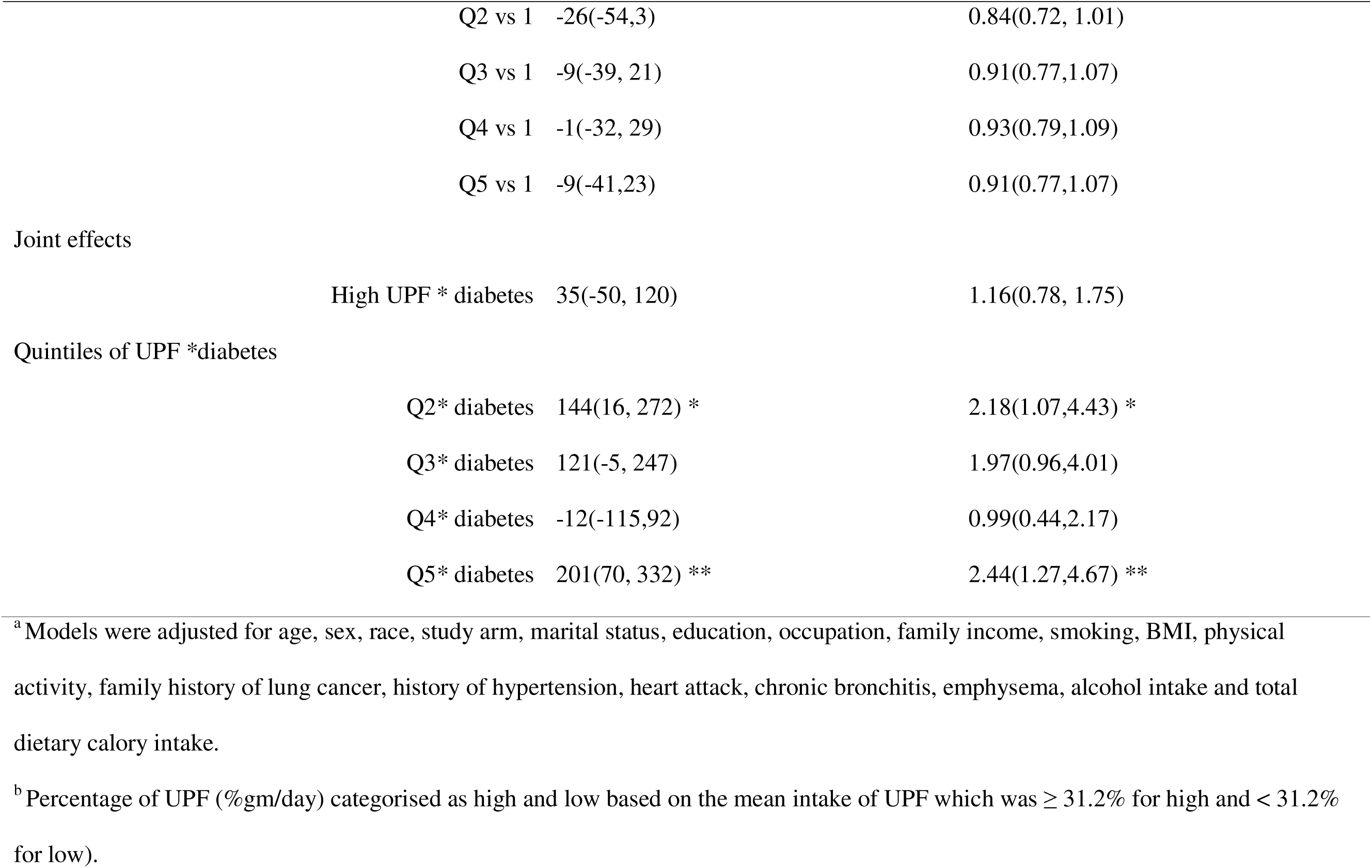
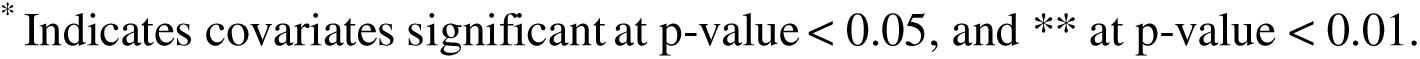
Interaction between UPF consumption and diabetes for the risk of lung cancer among adults using additive hazard models in comparison with Cox proportional hazard models, US (n = 91,453).

Sensitivity analyses were undertaken by considering different scenarios. First, we undertook multiple imputation for observations with missing values. The associations remained unchanged (i.e., similar with the complete-cases results). The second scenario was used to rule out reverse causation by excluding participants with diseases at baseline such as hypertension, diabetes, obesity, and LC cases that occurred during the first five years of follow-up. The results generally unchanged but participants in the highest quintile of UPF intake compared to the lowest had a 5% of higher risk of LC incidence when LC cases occurred in the first five years of follow-up were excluded (HR = 1.05; 95% CI: 0.83, 1.32; p-trend = 0.047) (**Supplementary Table 3**). The association did not also differ by the histopathological subtypes of LC. However, a 10% increase in the UPF intake (in % kcal/day) increased risk of LC by 32% (HR = 1.32; 1.15,1.52), a linear dose-response relationship (p-nonlinear = 0.82) (**Supplementary Table 3, Supplementary** Fig 4). Furthermore, when adjusted for artificial sweetening beverages, consumption of UPF was significantly increased the risk of LC by 53% (HR =1.53; 95% CI: 1.26, 1.86; p-trend =0.032). However, the risk of lung cancer was decreased by 2% among participants in the highest quintile of UPF intake compared to lowest when adjusted for dietary fiber intake (HR = 0.98; 95% CI: 0.83, 1.15; p-trend =0.864) (**Supplementary Table 3)**.

## Discussion

In this multi-center prospective study, we investigated the association between UPF intake in percentage weight (%g/day) and the risk of LC incidence among adults participating in PLCO trial in the US. The finding of this study did not demonstrate link between a higher proportion of UPF in the total diet (%g/day) and risk of LC across all participants. However, during subgroup analysis, the study showed that the risk of LC was twice as high among adults in the highest quintile of UPF intake compared to study participants without diabetes. Further analysis using Cox proportional model on additive scale showed the effect of interaction between UPF and diabetes is, in fact super-additive (where both RERI and AP are found to be greater than zero and SI is greater than one). Quantification of the public health burden attributed to the interaction between quintiles of UPF consumption and diabetes revealed a total of 201 additional incident LC cases per 100,000 person-years due to these joint effects. Additionally, the risk of LC was 21% higher in participants within the highest quintile of UPF intake according to total energy (% kcal/day) consumption compared to participants in the lowest quintile.

A few studies from EPIC, UK Biobank, and a meta-analysis have investigated a positive association between UPF consumption and overall cancer risk [11–13]. However, the association between UPF consumption and the risk of LC has not yet been well established, despite LC being the second most commonly diagnosed cancer globally, following breast cancer [40]. A national representative study from the UK demonstrated a borderline statistically non-significant higher risk (25%) of LC among people at the highest level of UPF intake compared to lowest [12]. In contrast, a study from European countries, EPIC study showed that there was a non-significant inverse association between UPF consumption and LC, with a 4% lower risk of LC among subjects at the highest level of UPF consumption [13].

The current study revealed a positive association between LC incidence and a higher consumption of UPF (% gm/day) only among participants who had diabetes at baseline. When UPF consumption was measured in terms of energy ratio (% kcal/day), the risk of LC increased among participants with highest level of UPF consumption.

The underlying mechanism for the observed association may be explained by the interplay between hyperglycaemia and central obesity-cancer linkage, resulted from insulin-resistance and insulin-like growth factor 1, and adipokine pathophysiology and systemic inflammation from circulating pro-inflammatory cytokines interleukin-6,8 and 1β, tumour necrosis factor-α, vascular endothelial growth factor, chemokine, ligand 2 and interferon [41–43]. A growing body of evidence supports the idea that abdominal obesity, which can be measured by waist circumference or waist-to-hip ratio, and metabolic dysregulation, such as hyperglycaemia, insulin resistance, and dyslipidaemia, may the underlying biological mechanisms that explain the link between consumption of UPF and the risk of LC. National representative prospective studies from Korea, the USA and the UK have emphasized the significance of considering metabolic status and markers when it comes to the primary prevention of lung cancer and the identification of high-risk populations for lung cancer screening [44–46].

The direct association of UPF consumption and risk of obesity, type 2 diabetes, and CVD are sufficiently well documented [8, 9, 47] and type 2 diabetes increases the risk of LC [48–50]. Studies showed that a 10 cm increase in waist circumference had 10% higher risk of LC [51, 52]. Central adiposity affects overall survival in LC and adiposity is inversely associated with inflammatory genes which may downregulate the anti-tumour immune response[53, 54]. Metabolic effect of central obesity encompasses deranged insulin signalling, increased steroid hormone signalling, increased glucose utilization, fatty acid utilization and aberrant adipokine signalling [55]. Generally, dietary patterns with higher UPF proportion are nutritionally inferior and high in energy, fat, hydrogenated fat, free sugars, and low dietary fiber, that all promote pro-inflammatory response, oxidative stress, metabolic dysregulation, and carcinogenicity [27, 44, 45]. This study highlighted that the observed association may not be affected by residual confounding, mainly by a history of cigarette smoking status, intensity, pipe, and cigar smoking. No significant subgroup difference was detected between UPF intake and risk of lung cancer by smoking status, BMI, family history of lung cancer and subtypes of lung cancer.

Moreover, during food processing alteration in food matrices contribute to degradation of essential nutrients involved in promoting health and change in microbiota [12, 21, 56]. Though evidence remains inconclusive, the mechanism for lung carcinogenesis in relation to UPF intake may be implicated due to metabolic disorders such as hyperglycaemia, insulin-resistance, dyslipidaemia, and inflammation due to altered levels of insulin-like growth factors, adipokines, myokines and sex hormones [57–59].

This study also determined whether the observed association between UPF intake (in %g/day, and %kcal/day and the risk of LC is altered by certain confounders such as age, sex, details of smoking status, baseline comorbidity, BMI, alcohol consumption, histological subtype, follow-up length, dietary risk factors such as fruits and vegetables, processed meat, artificial sweetening beverages, fish, fiber, sodium, total fats and other nutrients not largely changed. The risk of LC was higher among participants with NSCLC subtype and after excluding LC cases developed during the first five years of follow-up with level of UPF consumption increases. Adjusting for dietary fiber reduced the risk of LC, non-statistically significant while adjusting for artificial sweetening beverages significantly increased the risk. The association was not largely changed for other covariates.

Our finding using the weight contribution of UPF consumption showed no significant association with the risk of LC across all study participants, contrary to the association between caloric contribution and LC. This could be due to fact that the energy contribution of UPF can directly impact metabolic regulations and inducing oxidative stress than the weight contribution of UPF which comprises non-energy yielding constituents such as water, and artificial sweeteners [13, 33, 44, 45]. A well-designed, longitudinal study with adequate number of cases that incorporate biomarkers for central obesity, oxidative stress, inflammation, and additives and other non-nutritional factors, may help in reaching a definitive conclusion and provide a greater understanding of the biological mechanism(s) underlying the UPF consumption and LC association.

This study has important limitations. The NOVA grouping was performed using a single dietary data up to 25 years ago as a measure for usual dietary exposure. Since then, foods in the market have been continuously changed and UPF has become more dominate than minimally or unprocessed food groups in the American food system. Thus, the caloric and weight contribution of UPF may be underestimated and could not represent the current UPF consumption pattern in America. Recall bias from dietary assessment and misclassification bias from categorization of food items could not be ruled out even if a validated FFQ and NOVA food classification were used. The findings of the current study could only be inferred for the general aged 55 years and over and the follow-up duration is relatively short (12 years). The study did not investigate potential biomarkers that may mediate the observed association between UPF and LC such as measures of body fat composition, inflammatory factors and non-nutritional factors associated with UPF consumption (additives, preservatives and neoformed contaminants) and potential confounding factors related to environmental or occupational exposures.

## Conclusions

In summary, our findings showed that adults with diabetes and a higher consumption of UPF had a disproportionately higher risk of LC. In addition, a higher caloric contribution of UPF was positively associated with a higher risk of LC among all study participants. Given the accumulating evidence relating to the adverse effect of UPF intake on overall cancer risk, consideration of the degree of food processing and integrating public health policies that promote healthy foods may be critical in reducing the modifiable burden of LC. Limiting a higher level of UPF consumption and regulation of metabolic biomarkers could prevent a higher number of LC cases. Additionally, further research is needed to better understand the relationship between UPF consumption and the incidence of LC.

## Data Availability

All data produced in the present study are available upon reasonable request to the authors

## Acknowledgements

TCM is grateful to thank for support provided by the Australian Government Research Training Program Scholarship. The authors thank the National Cancer Institute for access to NCI’s data collected by the Prostate, Lung, Colorectal and Ovarian (PLCO) Cancer Screening Trial.

## Statement of authors’ contributions to manuscript

T.C.M: involved in conceptualization, data curation, formal analysis, investigation, methodology, project administration, resources, software, validation, visualization, writing the original draft, review & editing of the manuscript. Y.A.M, Z.S and T.K.G involved in conceptualization; methodology, supervision, validation, visualization, review and editing of the final version of the manuscript. All authors have read and approved the final manuscript. Data Sharing. The PLCO trial data are available upon request to The National Cancer Institute at <u>Access to PLCO Data, Images, and Biospecimens - Learn - PLCO - The Cancer</u> <u>Data Access System</u>.

## Abbreviations

BMI: Body Mass Index

BQ: Baseline Questionnaire

EPIC: European Prospective Investigation into Cancer and Nutrition

CVD: Cardiovascular disease

DHQ: Dietary History Questionnaire

FFQ: Food Frequency Questionnaire

LC: Lung Cancer

NCDs: Non-communicable Diseases

NCI: National Cancer Institute

PLCO: Prostate, Lung, Colorectal and Ovarian Cancer

UPF: Ultra-processed Food

UK: United Kingdom.

## Figure Legends

**Figure 1:** The relative contribution of food groups to the total diet (a) and to the subtotal of UPF consumption in men (b) and women (c) in the PLCO trial data.

**Figure 2:** Non-linear relationship of UPF consumption in weight ratio and lung cancer incidence among older adults participated in the PLCO trial, USA.

**Figure 3:** Subgroup analysis for the association between UPF consumption and risk of lung cancer incident stratified by demographic, lifestyle and baseline comorbidities in older adults enrolled in the PLCO trial, the in USA]. The model was adjusted for covariates mentioned in Table 2 footnote. BMI: body mass index; HR: hazard ratio; UPF: ultra-processed foods.

